# COVID 19 Pandemic: A Real-time Forecasts & Prediction of Confirmed Cases, Active Cases using the ARIMA model & Public Health in West Bengal, India

**DOI:** 10.1101/2020.06.06.20124180

**Authors:** Dibash Sarkar, Moinak Biswas

## Abstract

**Background:** COVID-19 is an emerging infectious disease which has been declared a Pandemic by the World Health Organization (WHO) on March 11 2020. This pandemic has spread over the world in more than 200 countries. India is also adversely affected by this pandemic, and there are no signs of slowing down of the virus in coming time. The absence of a vaccine for COVID-19 is making the situation worse for the already overstretched Indian public health care system. As economic burden makes it increasingly difficult for our country to continue imposing control measures, it is vital for states like West Bengal to make predictions using time series forecasting for the upcoming cases, test kits, health care and estimated the requirement of Quarantine centers, isolation beds, ICU beds and ventilators for COVID-19 patients.

**Objective:** This study is forecasting the confirmed and active cases for COVID-19 until August, using time series ARIMA model & Public Health in West Bengal, India.

**Methods:** We used ARIMA model, and Auto ARIMA model for forecasting confirmed and active cases till the end of August month using time series data of COVID-19 cases in West Bengal, India from March 1, 2020, to June 4, 2020.

**Results:** We are expecting that West Bengal will have around **62279 ± 5000** Cases by the end of August based on our forecasts. Meanwhile Maharashtra, Punjab, Gujarat and Delhi (UT) will be the most affected states, having the highest number of active and confirmed cases at the end of August.

**Discussion and Conclusion:** This forecasts show a very crucial situation for West Bengal in coming days and, the actual numbers can go higher than our estimates of confirmed case as Lockdown 5.0 & Unlock 1.0 will be implemented from 1^st^ June 2020 in India, West Bengal will be observing a partial lift of the lockdown and in that case, there will be a surge in the number of daily confirmed and active cases. The requirement of Health care sector needs to be further improved isolation beds, ICUs and ventilators will also be needed to increase in that scenario. Inter State & Intra State Movement restrictions are lifted. Hence, Migrants returning to their homes due to loss of livelihood and income in the lockdown period may lead to a rise in the number of cases, which could not be accounted for in our projections. We suggest more of Public-Private Partnership (PPP) model in the health sector to accommodate COVID-19 patients adequately and reduce the burden of the already overstretched Indian public health care system, which will directly or indirectly affect the States in the time of crisis.

## Introduction

Coronavirus disease (COVID-19) is one of the greatest challenges the world has encountered in recent times. Since the initial reports of outbreak in late December, 2019, It is caused by severe acute respiratory syndrome Corona Virus 2 (SARS-CoV-2). The origin of the virus is yet to be confirmed, but the first person tested positive is from Wuhan, China. It is spreading very quickly throughout the world & the numbers have been consistently rising with the disease affecting 6.54 million people in 181 countries worldwide as of 6^th^ June, 2020. In India, the first positive case of COVID-19 was detected on January 30^th^ 2020, in Kerala. The frailty of a multitude of health care systems across the globe has been exposed by COVID-19. With the surfacing negative socioeconomic consequences of community mitigation strategies like lockdown affecting the vulnerable, especially in developing countries like India, Governments are eyeing at easing the restrictions that have long been in place by recent Lockdown 5.0. It is imperative to understand that lifting the control measures for economic salvage, without thoroughly preparing for the possible consequences, may only result in further economic decline and health crisis.

WHO in its strategic advice for countries looking to life the control measures illustrated six criteria in a sequential manner to be considered: control of transmission; preparation of health systems for active contact tracing and optimum care provision; careful management of health facilities to prevent outbreaks; adherence to preventive measures as the essential services resume; management of importation risks; indoctrination of the ‘new norm’ among communities by active engagement. According to the report of World Population prospects (2019), India has a population of more than 1.36 billion and most of the population of urban areas and cities are under the risk of contracting the virus. So, it is important to forecast numbers of confirmed and active cases. In this scenario, it is vital for every state to make predictions using time series forecasting, the study aims to draw comparative account of the progression of COVID-19 in near future in the state of West Bengal.

### Data & Methodology

#### Data

For our study, the required data of daily total confirmed cases and total active cases of COVID-19 infection collected for India as well as its state West Bengal from the (https://www.covid19india.org/), and excel of the patient database is used to build a time-series database for confirmed and active cases Using the Data Science Software called R. In this study, forecasting is done based on the data from March 1, 2020, to June 4 2020. This data is being used to build Forecast which includes all the Statistics and Graphs for mentioned model.

## Methodology

In the past few months, an increasing number of research related to forecasting the trend of pandemic COVID-19 cases are being published using different approaches in various part of the world. (Gupta and Pal 2020; Fanelli and Piazza 2020; Giordano et al. 2020; Tandon et al., 2020; Kumar et al. 2020; Benvenuto et al. 2020; Batista, 2020). The ARIMA model is one of them and nowadays used for forecasting case count for the prediction of epidemic diseases based on the time series modeling (Rios et al. 2000; Li et al. 2012; Zhang et al. 2014; Benvenuto et al., 2020).

In this study, the well-known Autoregressive Integrated Moving Average (ARIMA) time-series model used for the further forecasting purpose. ARIMA model is one of the generalized forms of an autoregressive moving average (ARMA) model among the time series forecasting. We fit both models to understand the data better or to predict future points in the series (Forecasting). ARIMA model depends or always represented with the help of some parameters, and the model has expressed as ARIMA (p, d, q): p, d and q are non-negative integers.

The parameters have their usual meaning, here, *p* stands for the order of auto-regression, *d* represents the degree of trend difference (the number of times the data have had past values subtracted) for the stationary of the trend and *q* signifies the order of moving average. This model combines auto regression lags under the stationary trend and moving average and predict better future values based on past and recent data. For this model, the degree of parameters *p*, d and q determine based on the partial Auto-correlation function (PACF) graph, The Augmented Dickey-Fuller Test to test the stationary of the time series observations and Complete Auto-Correlation Function (ACF) graph respectively (Forecasting COVID-19 cases in India)

We have applied the ARIMA model and Auto ARIMA model using R, to our considered time series data of COVID-19 cases in West Bengal for the forecasting the total confirmed and active cases for West Bengal and its majorly affected Districts. We selected Districts based on the criterion that chosen Districts should have at least 2 confirmed cases till June 2^nd^ 2020. By using this selection criterion, West Bengal and its 24 Districts Alipurduar, Bankura, Birbhum, Cooch Behar, Dakshin Dinajpur (South Dinajpur), Darjeeling, Hooghly, Howrah, Jalpaiguri, Jhargram, Kalimpong, Kolkata, Malda, Murshidabad, Nadia, North 24 Parganas, Paschim Medinipur (West Medinipur), Paschim (West) Burdwan (Bardhaman), Purba Burdwan (Bardhaman), Purba Medinipur (East Medinipur), Purulia, South 24 Parganas, Uttar Dinajpur (North Dinajpur), The cases are forecasted under the assumption that people will be maintaining condition similar to the partial lockdown situation and maintain physical distancing with self quarantine. After fitting the model, the built model is used to forecast confirmed and active cases COVID-19 cases for the next 80 days, *i*.*e*. from June 6^rd^, 2020, to August 3^rd^ 2020.

The model for forecasting future confirmed and active cases of COVID-19 cases is represented as,

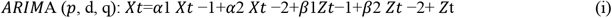

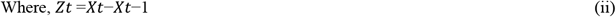

Here, *Xt* is the predicted number of confirmed and active COVID-19 cases at *t*^*th*^ day; *α*1, *α*2, *β*1 and *β*2 are parameters whereas *Zt* is the residual term for *t*^*th*^ day.

The trend of forthcoming incidences can be estimated from the previous cases, and a time series analysis is performed for this purpose (Tandon et. al., 2020).

In our study, the forecasted cases are mainly used for preparing the government of West Bengal for the health infrastructure such as the number of isolation beds, ICU beds and ventilators, Quarantine centers etc. across the State. In further analysis, based on predicted active cases, we estimated the hike in number of cases by COVID-19 patients in the coming days. Based on this theory we can maximize the requirement of Quarantine centers, Alcohol based Sanitizers, Public sanitizing materials, ICUs and ventilators as the infection hit its peak, which the state may get in the month of July-August according to the forecast.

Our health infrastructure requirement is estimated based on the active cases as our projections are made on the basis on data till June 2^nd^ when our country was observing the complete lockdown and starting of Lookdown 5.0. However, India is observing partial lockdown in Containment Zones currently and has removed lockdown in the Red, Yellow & Green Zones, so for being prudent, we will estimate the amount of active cases which are to be increased and also look into the Public Health Infrastructure of West Bengal.

## Results

### Forecast of confirmed and active cases by the ARIMA model for West Bengal

**Correlogram and ARIMA forecast graph for the Confirmed COVID-19 Cases in West Bengal**

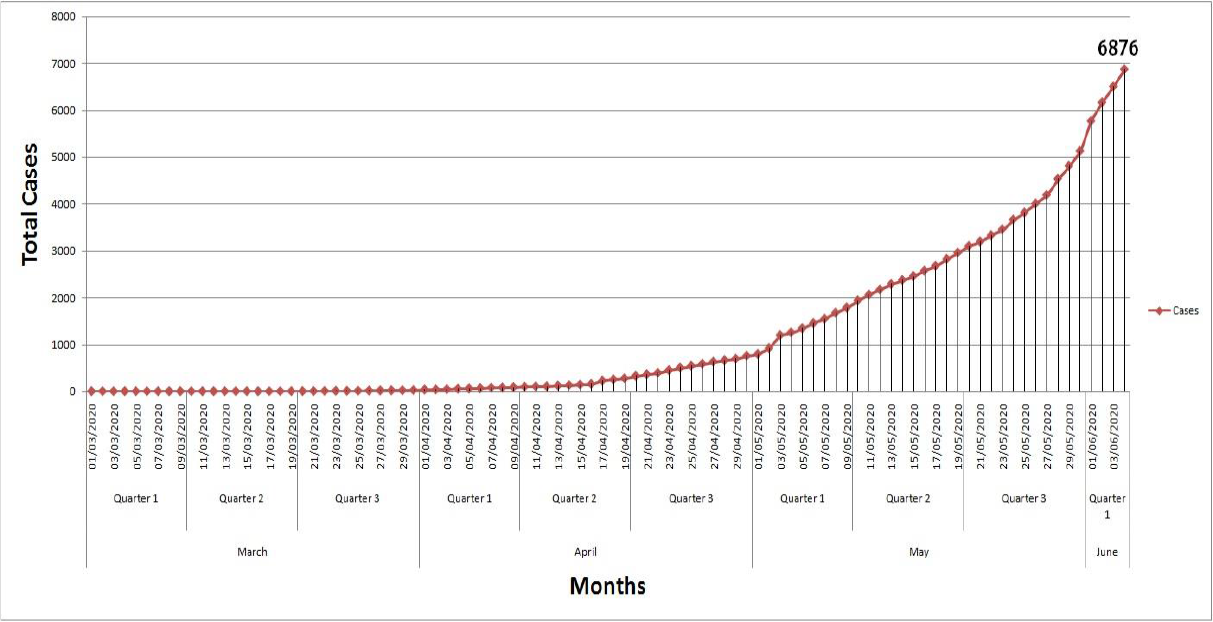

**Fig. – Correlogram of Total COVID - 19 Cases**.

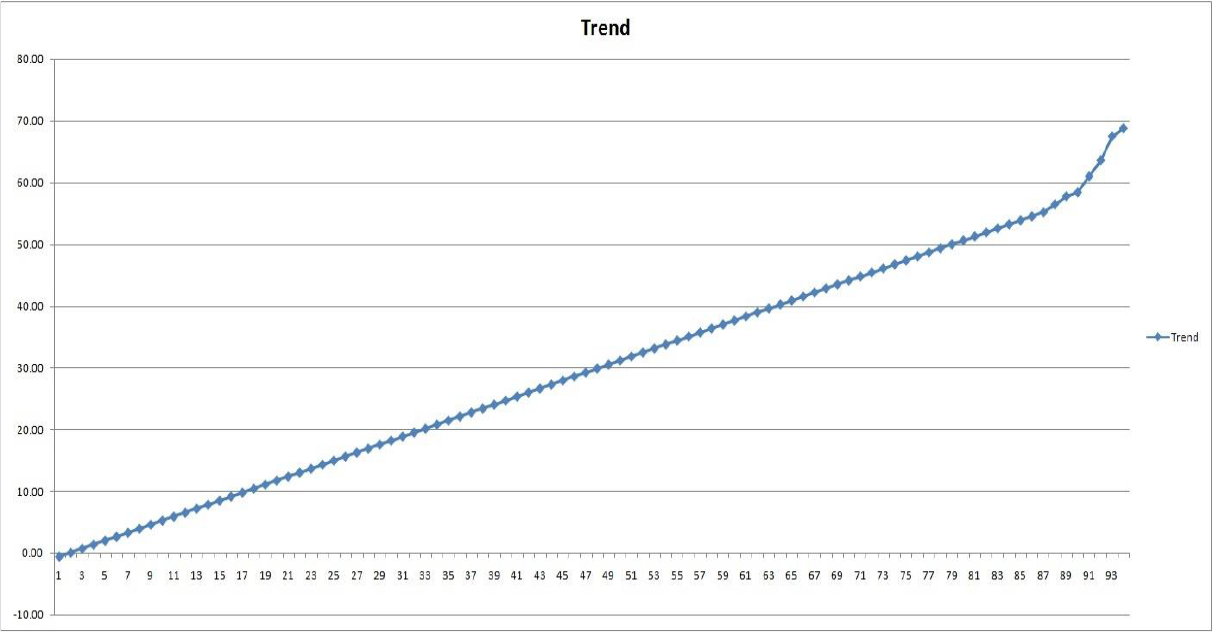

**Fig. – Trend of COVID - 19 Cases in West Bengal**.

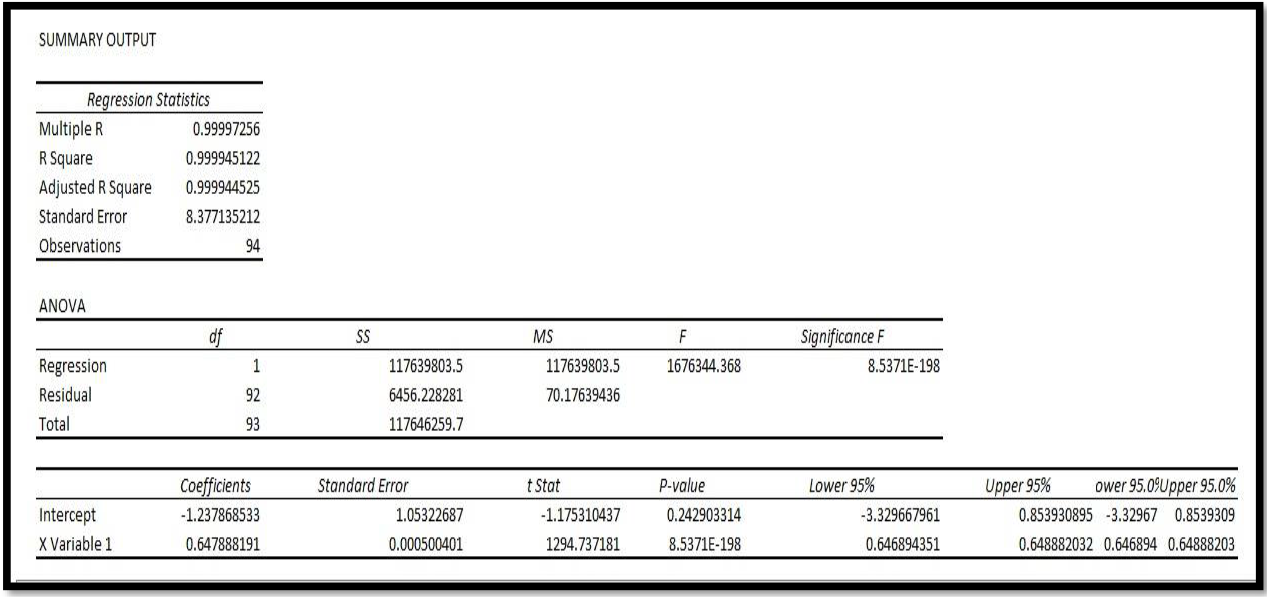

**Fig. - Regression Table for the Total Data**

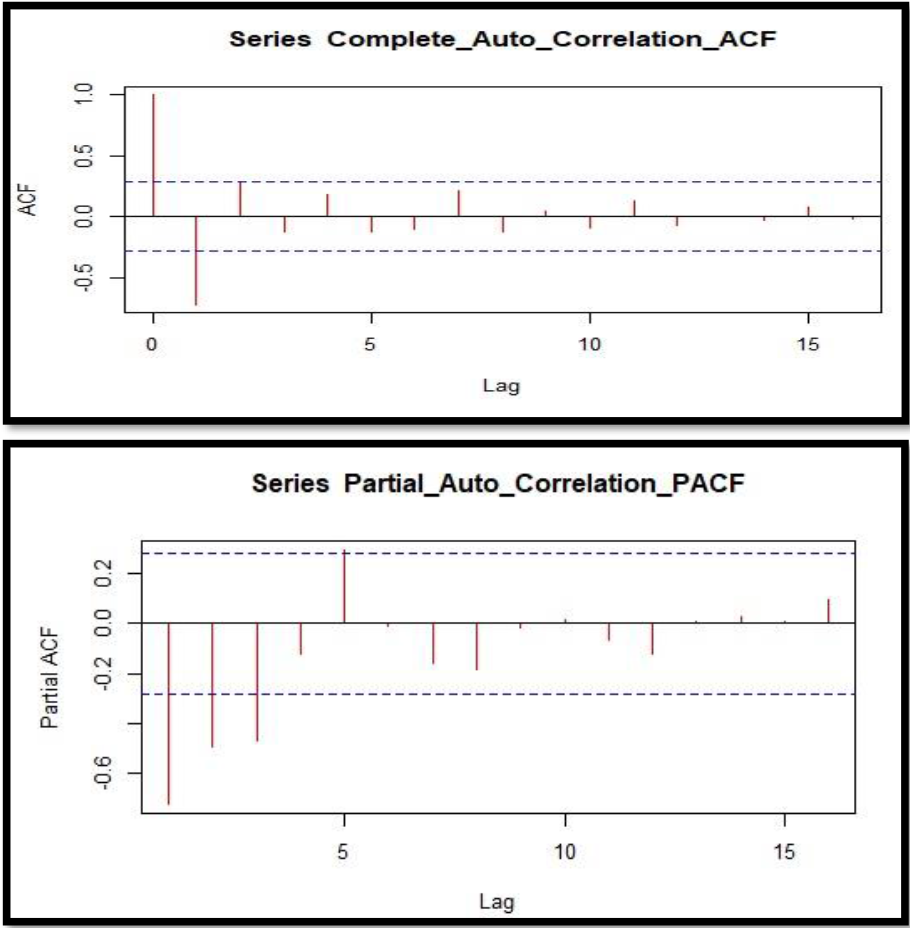

**Fig. - The ACF and PACF plot for the determine the value of q and p for the model**.

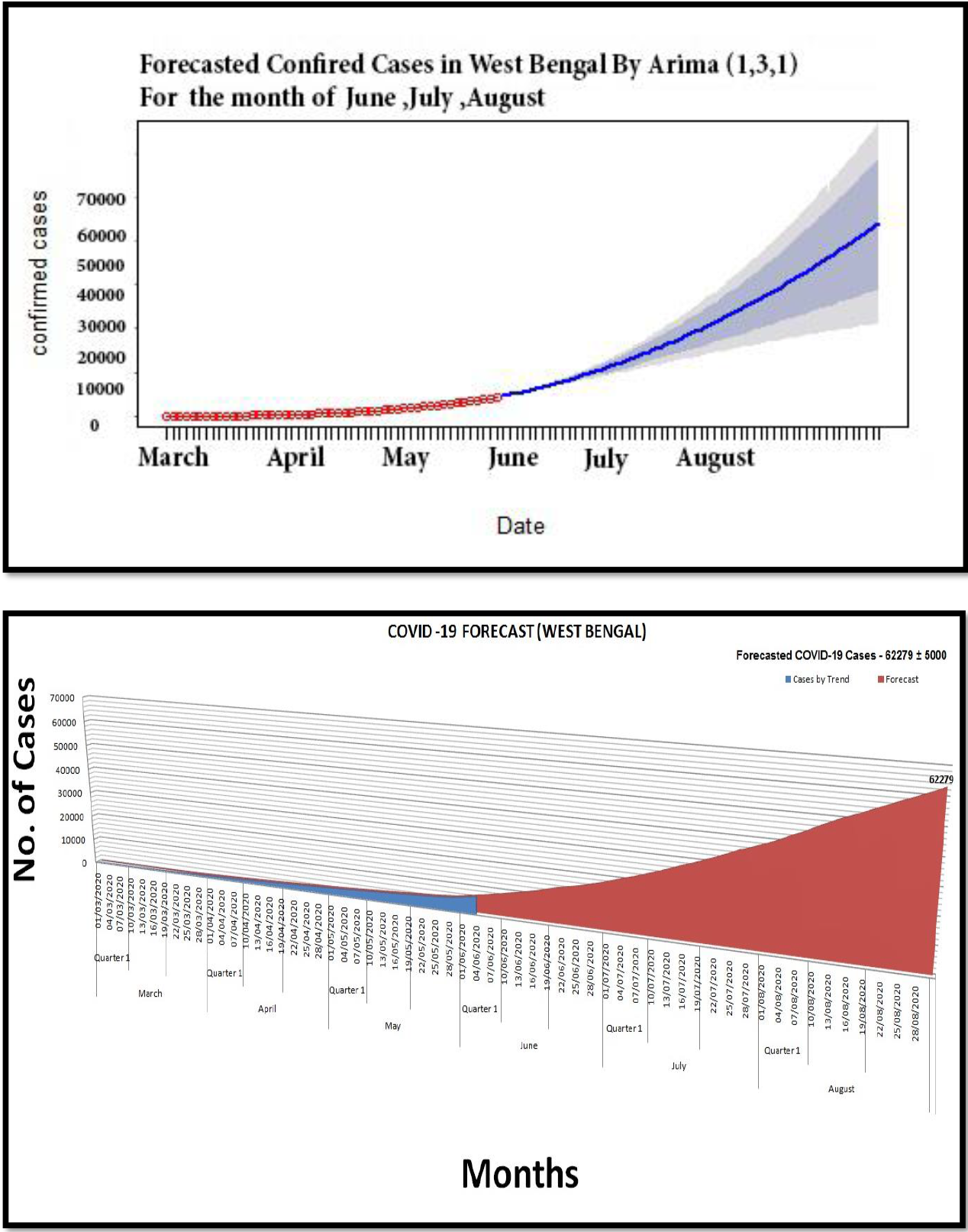

**Fig**.**- COVID – 19 Forecast For the month of July & August Estimated Cases – 62279 ± 5000**

### District - wise Confirmed cases data** in West Bengal

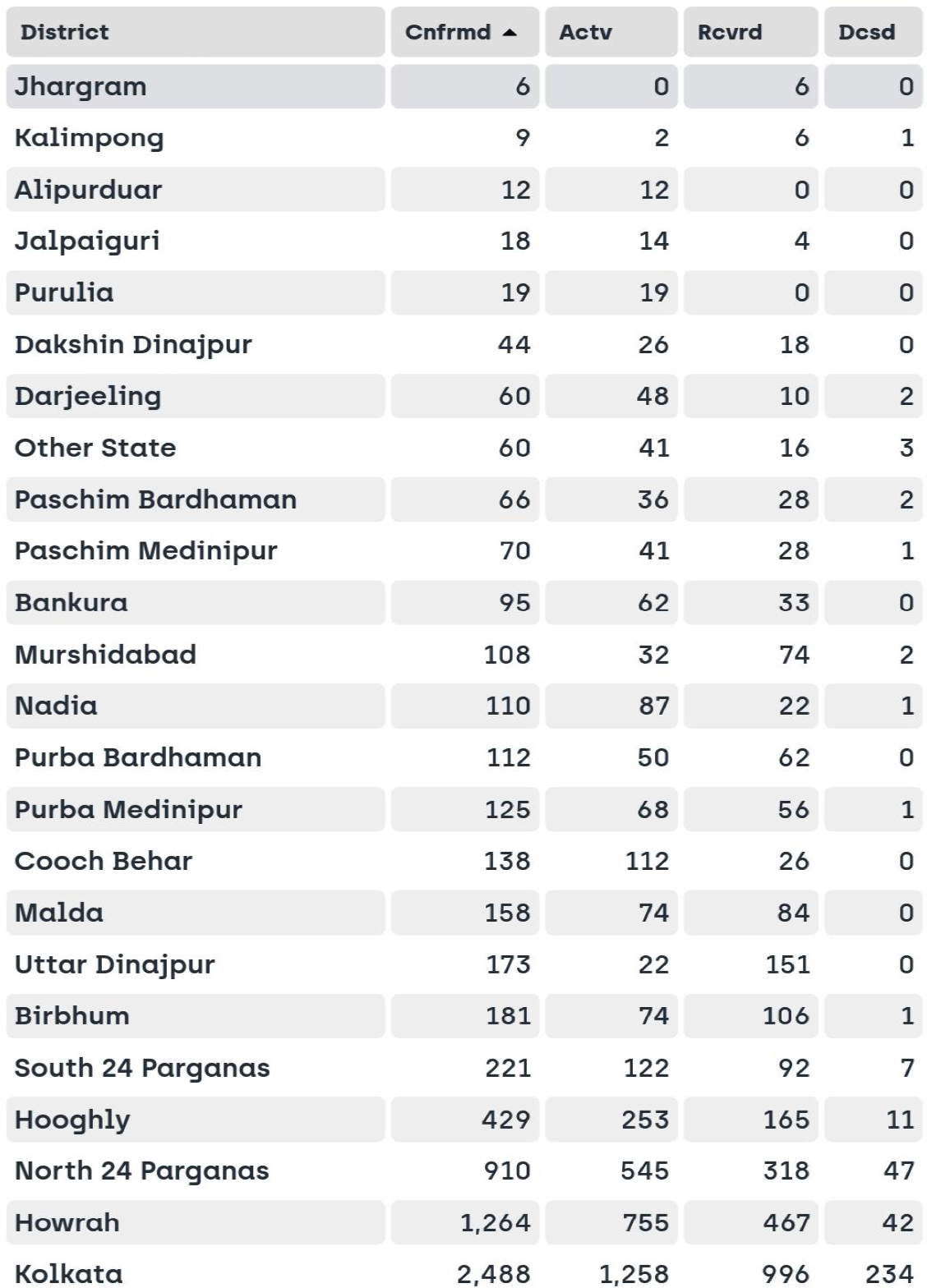

***\* * Data is taken from West Bengal Govt. COVID-19 Website (till 4th June 2020)***

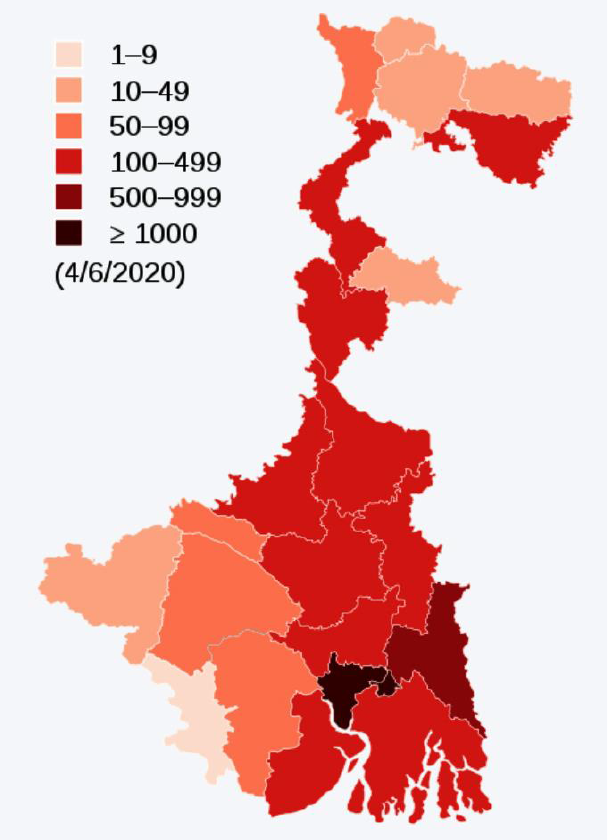

**Fig: Map of Districts with Confirmed Cases**

### Top Districts Affected With COVID-19 in West Bengal

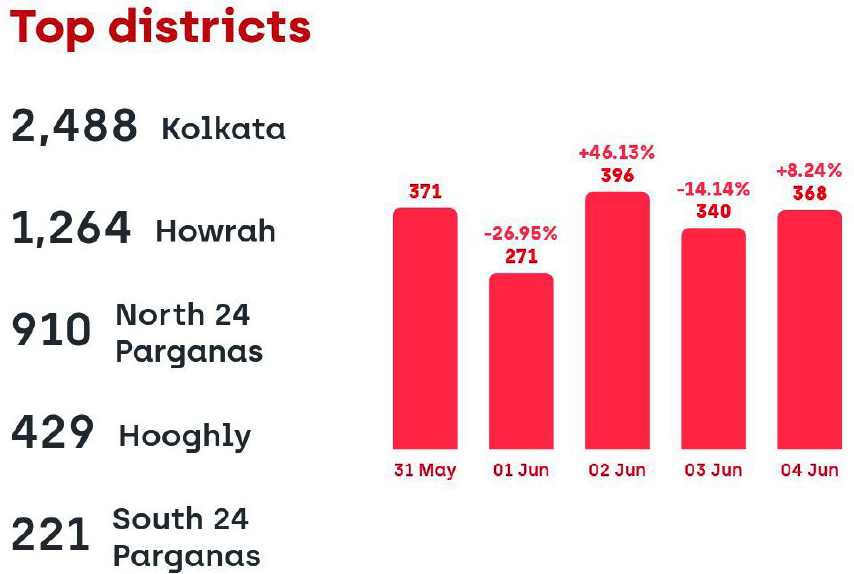

***\* * Data is taken from West Bengal Govt. COVID-19 Website (till 4th June 2020)***

### Kolkata’s Covid-19 Figure: A Case Study

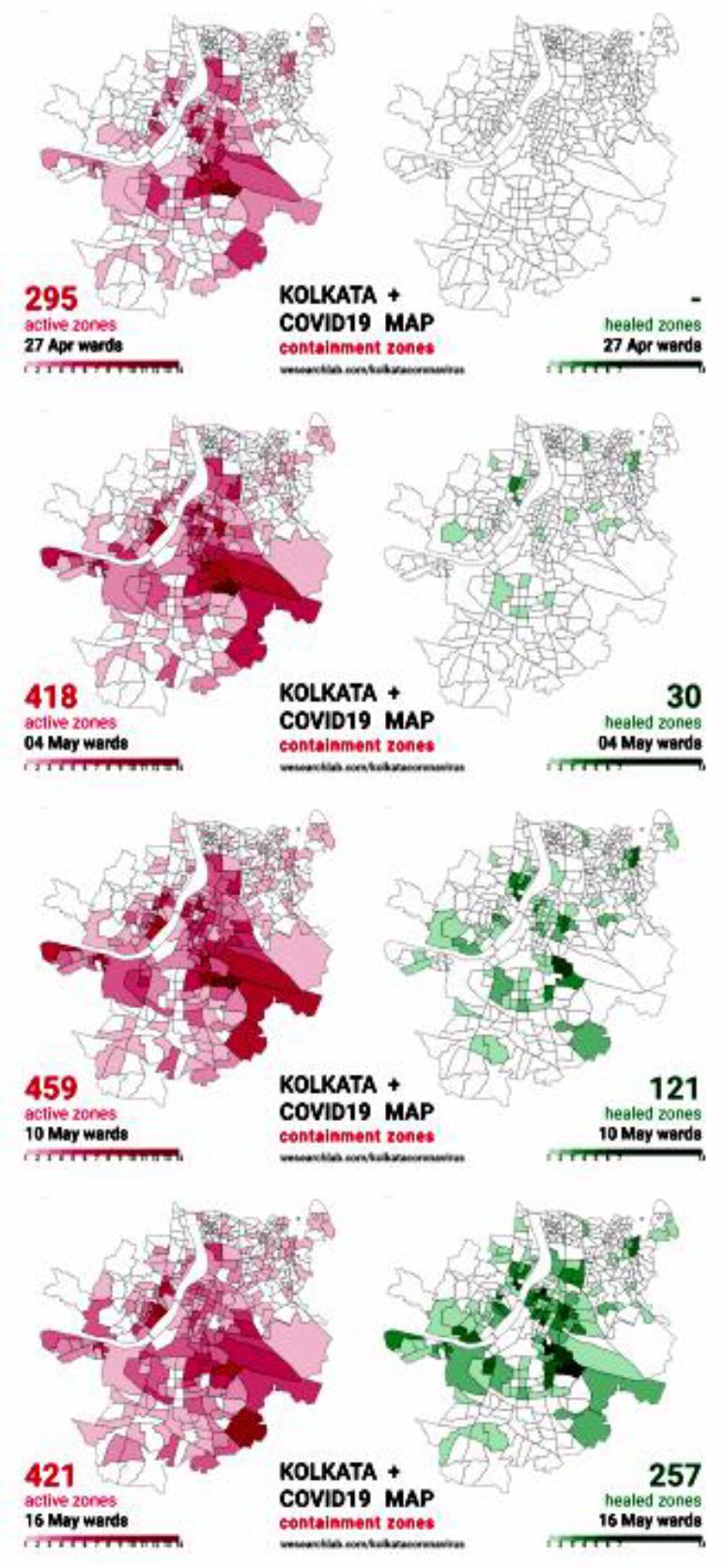

##### Kolkata’s COVID-19 containment zones

The centre has identified 170 COVID-19, hotspot districts in 25 states across India. The states have been asked to classify hotspot areas as red zones and focus on converting the red zones to orange and then green zones.

The West Bengal government on Monday released a list, saying four districts, including Kolkata, have been declared as red zones in the wake of the COVID-19 outbreak, and 348 areas as containment zones, out of which Kolkata has 227. There are some districts in Orange & Green zones. The lockdown process will be strict in those Red Zone Containment areas. Some relaxation will be given to Orange zones and most relaxation will be given to Green Zones.

The West Bengal government has been sharing regular updates of the lists of containment zones from 4^th^ May 2020.

### COVID-19 Pandemic Data in West Bengal

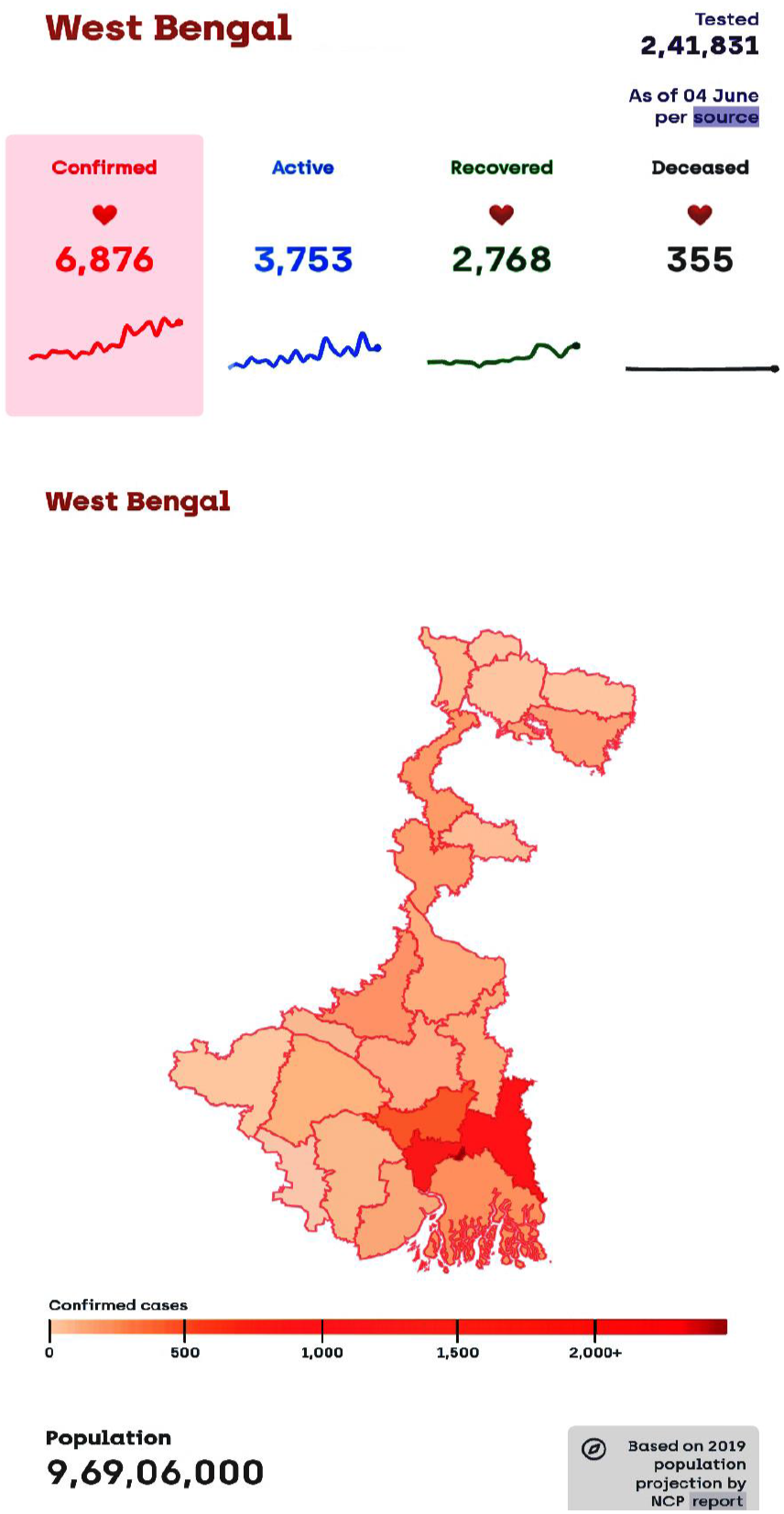

### Confirmed Per Million, Active, Recovery Rate, Mortality Rate, Average Growth Rate, Tests Per Million

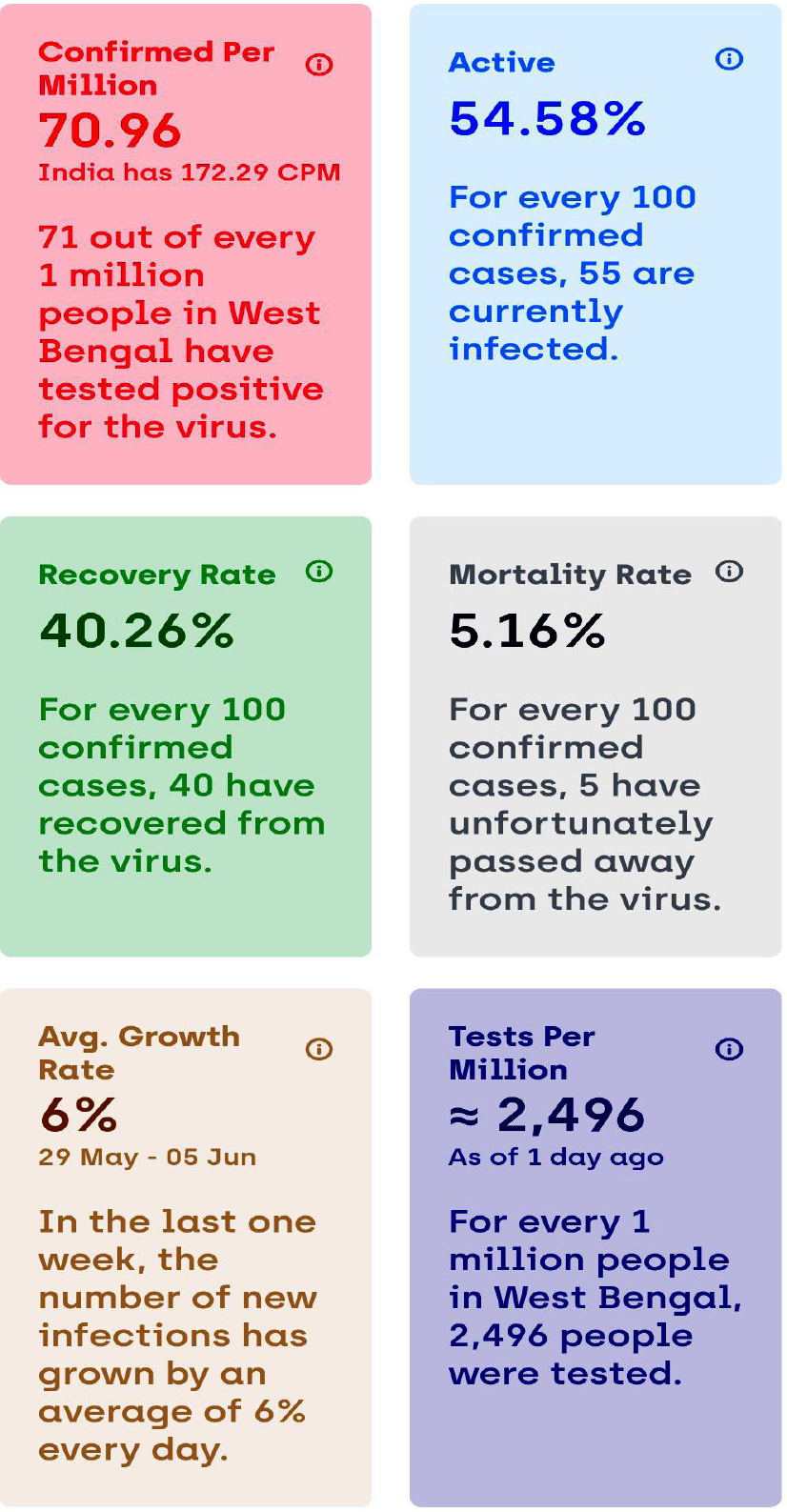

### Spread Trends of COVID-19 IN West Bengal

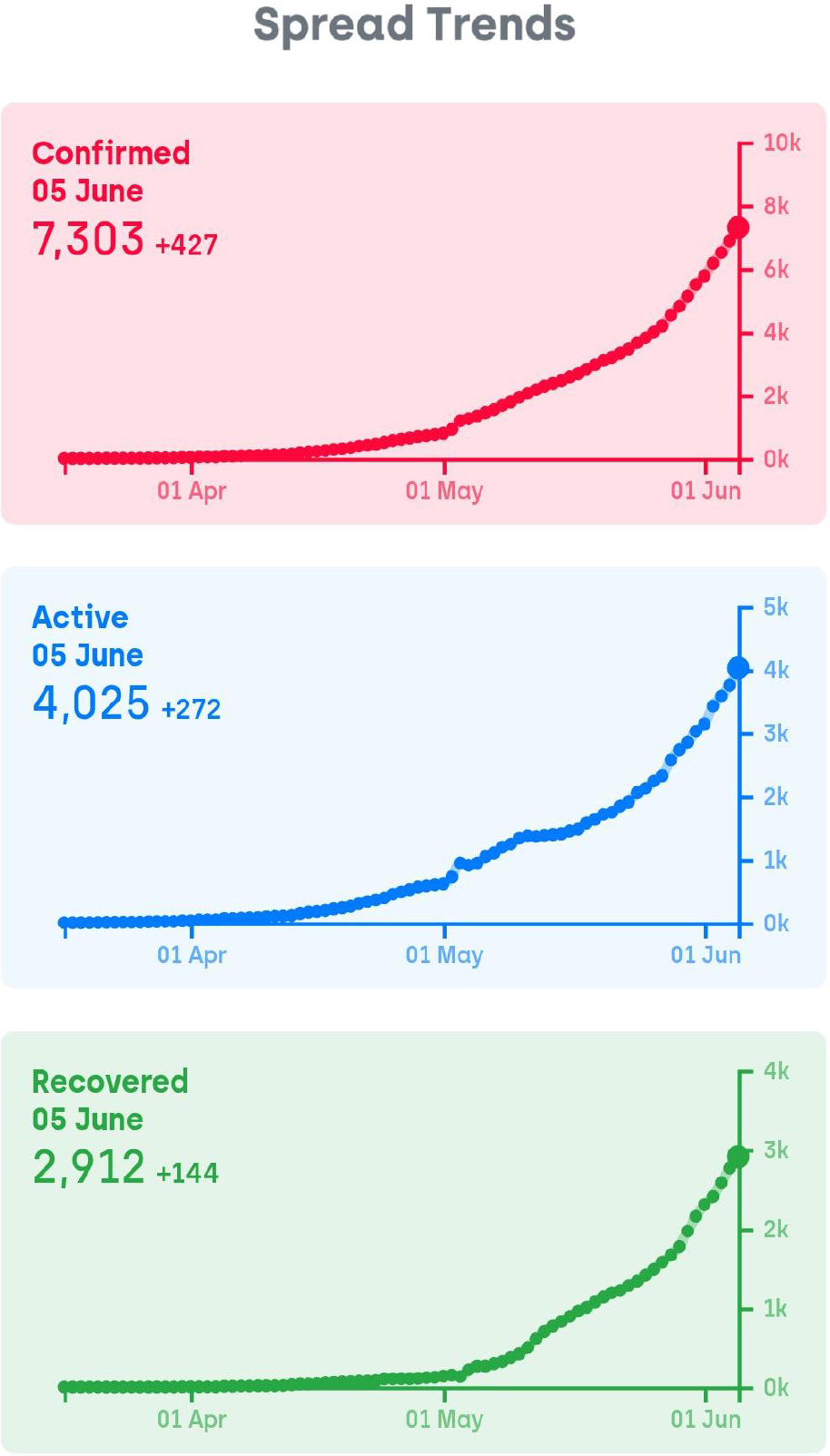

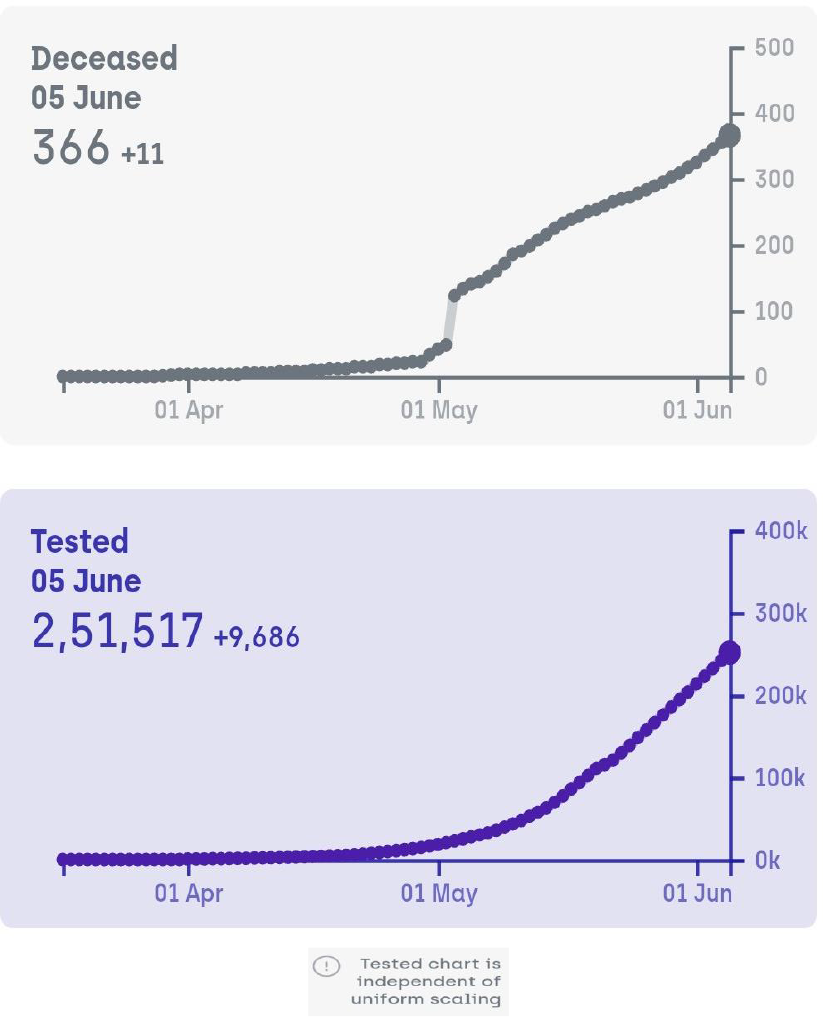

**Fig: Spread Trends of COVID-19 IN West Bengal**

### COVID-19 Cases** in INDIA

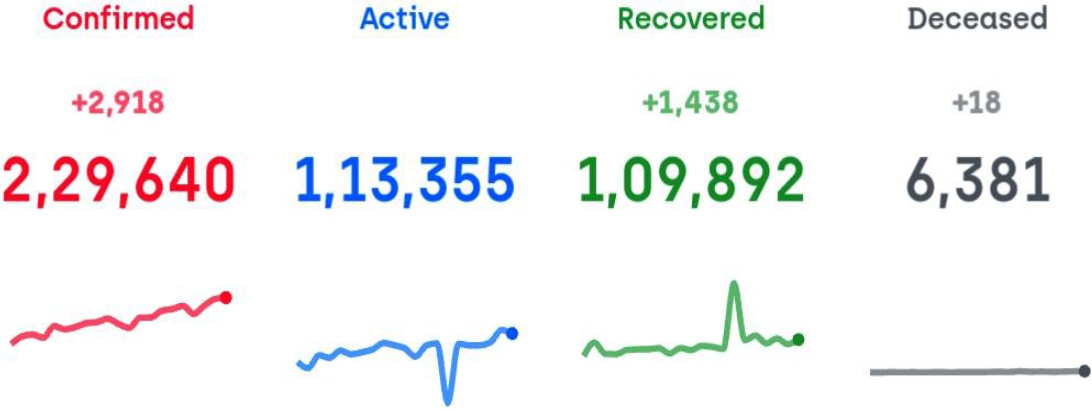

***\* * Data is taken from West Bengal Govt. COVID-19 Website (till 5th June 2020)***

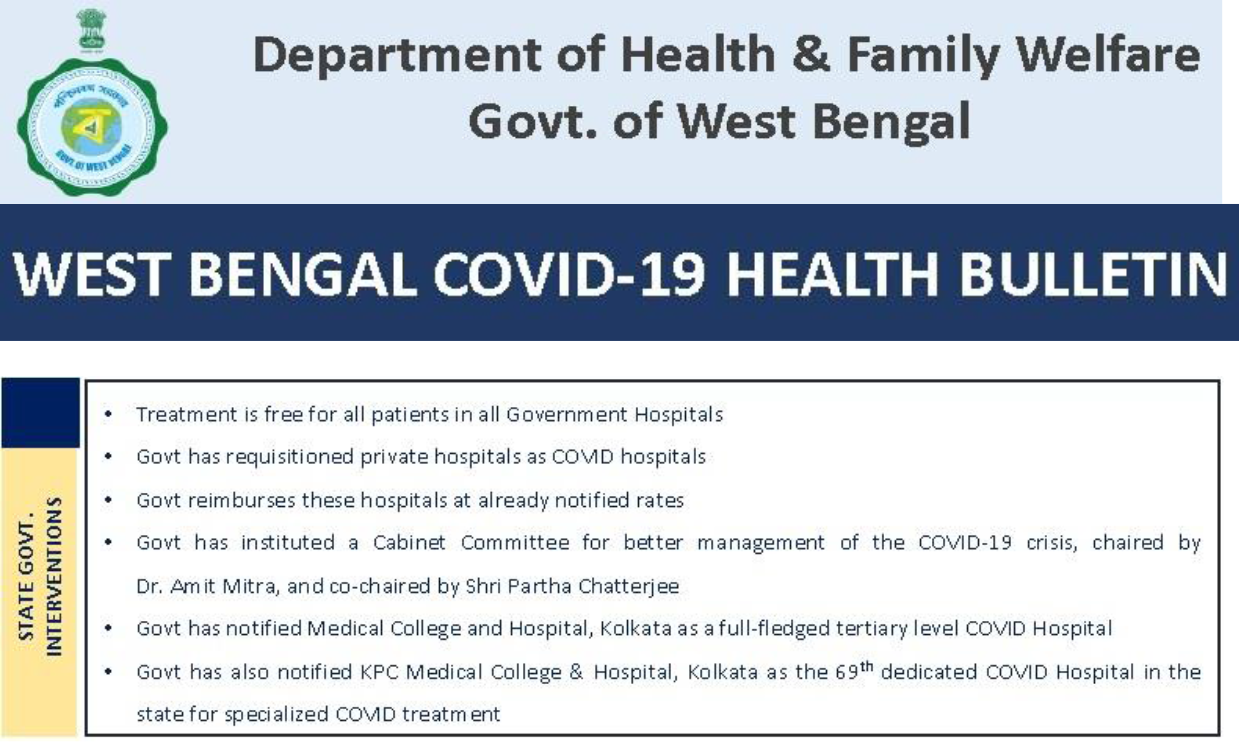

***Fig: State Govt. Interventions for COVID-19***.

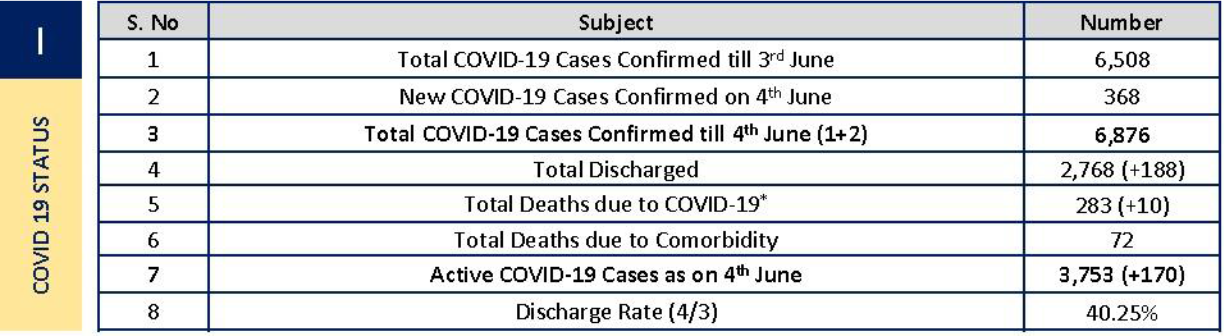

***Fig: COVID-19 Status in West Bengal***.

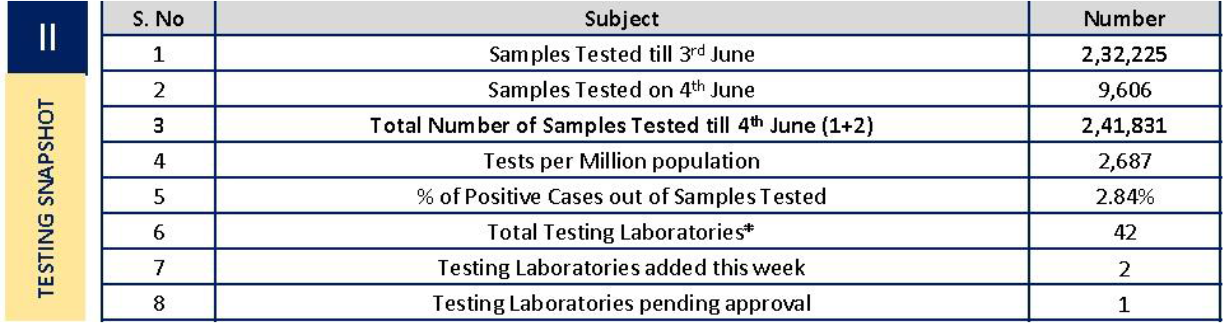

***Fig: Sample Testing Stats for COVID-19***.

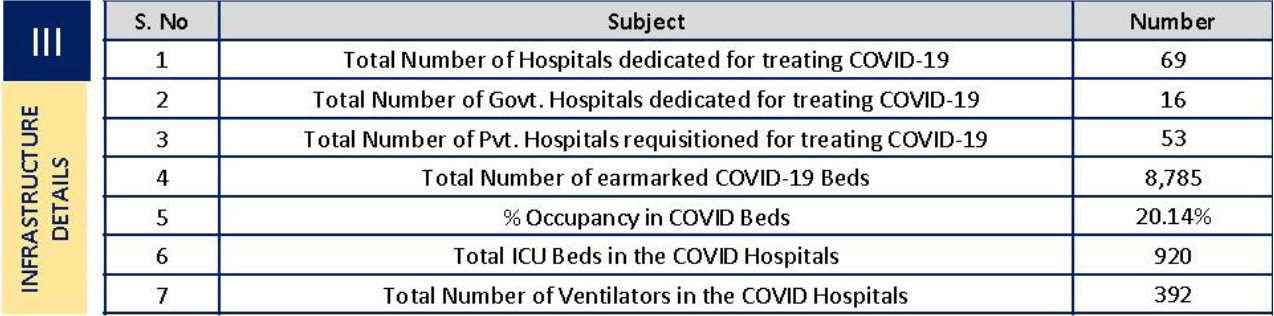

***Fig: Healthcare Infrastructure for COVID-19***.

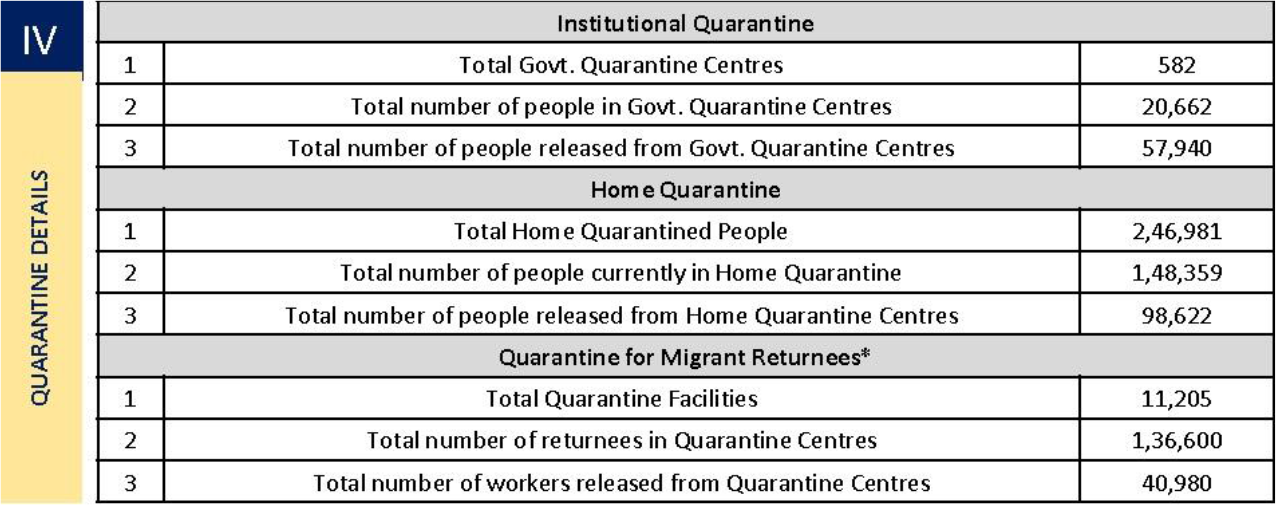

***Fig: Quarantine Details for COVID-19***.

### Allegations against West Bengal Government: A Case Study

Chief Minister of West Bengal Mamata Banerjee and her government was widely criticized of the handling of the coronavirus pandemic and was accused of concealing facts by the opposition and critics. The opposition accused Mamata of playing ‘appeasement politics’ amid the COVID-19 crisis. On 1 April, Banerjee claimed that the West Bengal Government have already traced 54 people who attended the Tablighi Jamaat religious gathering during the COVID-19 Outbreak, and 44 of them are foreigners. Although according to a report by central security agencies, 232 people had attended the Delhi’s Tablighi Jamaat event from West Bengal.

Of this, 123 are Indian nationals and 109 are foreigners.

Sooner she clarified that her government has acted swiftly after the Nizamuddin area was declared as a hotspot where nearly 2,300 people were staying despite the lockdown.

She further added that the government has quarantined 177 people, including 108 foreigners, who attended the Tablighi Jamaat congregation at the Nizamuddin Markaz.

The West Bengal Government has been also criticized for not sending enough samples to the National Institute of Cholera and Enteric Diseases(NICED) for testing.

West Bengal test numbers saw some rise after talks between government and NICED. According to them, this will be scaled up further in coming days.

The West Bengal Government has also been recommended to ensure transparency, genuine and verifiable data of COVID-19 by the West Bengal Doctors Forum (WBDF), as doctors cannot afford to send wrong signals to the world.

The doctors also hit out at the idea of the bureaucratic system to identify the death of COVID-19 patients. Their spokesperson claimed that every doctor is qualified enough and does not need a committee for such certification. On April 25, 2020,

The WB Govt admitted that 57 COVID-19 patients died but also said that 39 from comorbidities, after Inter Ministerial Central Team (IMCT) seek report.

The IMCT also pointed out flaws of the Govt in their letter to the Chief Secretary Rajiva Sinha, in which the letter read:

There were a large number of patients in the isolation wards of Chittaranjan National Cancer Institute (CNCI) as well as MR Bangur hospitals awaiting COVID test results for five days or longer.

Specifically at CNCI, there were four patients awaiting test result since April 16, 2020, two patients awaiting test result since April 17, 2020, and three since April 18, 2020.

Some of the patients have tested negative. It is not clear why the test results should take such a long time and there is a danger of COVID-19 negative patient acquiring the infection in the hospital while awaiting test result

## Discussion& Conclusion

The world is going through a pandemic, and almost every country is affected by it. A country as well as the States needs to know how much burden of active and confirmed cases it will have to bear in the coming time. It will help the states in taking pre-active measures to prepare adequate health infrastructure for the coming time based on future needs. We used ARIMA model and Auto ARIMA model on the time series data of COVID-19 cases in West Bengal for forecasting the total confirmed and active cases till August end.

Based on the forecasts, confirmed cases for West Bengal at the end of June are expected to be 17838-18724 (95% CI: 128806, 227968). West Bengal will be having 27147-30616 confirmed cases (95% CI: 173917, 415800) in the mid of July from the estimates, Even West Bengal will be having 50588-55617 confirmed cases (95% CI: 198917, 525800) in the mid of August from the estimates & we expect that India will be having

**Estimated Cases – 62279 ± 5000** at the end of August.

These results also show that daily confirmed cases are increasing at a faster pace even at the end of June with around 400-500 daily confirmed cases, so it is likely that peak will be attained afterwards.

According to our forecasts, it is a very alarming situation for India & West Bengal in coming days. However, the actual numbers can go higher than our estimates of confirmed cases, active cases & trends we made based on the data till June 3rd in this forecast, when West Bengal observed complete lockdown. Currently, West Bengal has a partial lockdown or Following Unlock 1.0 with restrictions varying for three zones (red, orange and green zone) based on the current assessment of the situation in there.

Lockdown is getting lifted, and in this case, there will be a surge in the number of daily confirmed and active cases.

The requirement of isolation beds, ICUs and ventilators will also be increased in that scenario. The migrants are returning to their homes due to loss of livelihood and income in the lockdown period, which may lead to a rise in the number of cases, and could not be accounted for, in our projections.

So, India and its majorly affected states like Maharashtra, Gujarat, Tamil Nadu and Delhi & West Bengal need to be well prepared for the pandemic challenge in coming time and focus on increasing their healthcare infrastructure, and other states should also remain alert till the pandemic completely recedes. We suggest a Public-Private Partnership (PPP) model in the health sector to accommodate COVID-19 patients adequately and reduce the burden of the already overstretched Indian public health care system.

### Limitations or Errors that may occur

The forecasting of COVID-19 cases is done based on the data under the lockdown duration and few in Unlock 1.0. So, the forecasted cases in future will be showing the same trend as India would have observed, had it been observing complete lockdown.

Since May 4, India is observing Unlock 1.0, and for that actual cases will/can/may be more than the forecasted cases. For our state it is showing hike in COVID-19 infection and increased trend in future, but the situation may not occur in future because of the nature of the previous trend-pattern is different from now.

Forecasted cases based on ARIMA model in our study for some states having lower bound for the 95% CI comes negative values which we have considered zero cases in that situation. In our study, the seasonality factor was considered but it may vary now due to Unlock 1.0, and it may affect our Forecast, Therefore the is a Plus – Minus in the forecasted cases to avoid any Big Error, & make the Data more Reliable.

**Table.** Date wise number confirmed and active cases in West Bengal, Trend, Predicted

| | | | $Y_t$ | | $Y_t/CMA$ | Average (respective quature) | De-Seasonalized Value | Intersept +Slope*Time Code | |
| --- | --- | --- | --- | --- | --- | --- | --- | --- | --- |
| No. | Months | Date | Cases | Moving Average (5) MA | $St * It$ | $St$ | $Y_t/St$ | Trend | Predicted ($Y_p$) |
| 1 | March | 01/03/2020 | 0 |  | 0.00 | 0.00 | 0.00 | -0.59 | 0 |
| 2 |  | 02/03/2020 | 0 | 0.00 | 0.00 | 0.00 | 0.00 | 0.06 | 0 |
| 3 |  | 03/03/2020 | 0 | 0.00 | 0.00 | 0.00 | 0.00 | 0.71 | 0 |
| 4 |  | 04/03/2020 | 0 | 0.00 | 0.00 | 0.00 | 0.00 | 1.35 | 1 |
| 5 |  | 05/03/2020 | 0 | 0.00 | 0.00 | 0.00 | 0.00 | 2.00 | 1 |
| 6 |  | 06/03/2020 | 0 | 0.00 | 0.00 | 0.00 | 0.00 | 2.65 | 2 |
| 7 |  | 07/03/2020 | 0 | 0.00 | 0.00 | 0.00 | 0.00 | 3.30 | 3 |
| 8 |  | 08/03/2020 | 0 | 0.00 | 0.00 | 0.00 | 0.00 | 3.95 | 4 |
| 9 |  | 09/03/2020 | 0 | 0.00 | 0.00 | 0.00 | 0.00 | 4.59 | 5 |
| 10 |  | 10/03/2020 | 0 | 0.00 | 0.00 | 0.00 | 0.00 | 5.24 | 6 |
| 11 |  | 11/03/2020 | 0 | 0.00 | 0.00 | 0.00 | 0.00 | 5.89 | 7 |
| 12 |  | 12/03/2020 | 0 | 0.00 | 0.00 | 0.00 | 0.00 | 6.54 | 9 |
| 13 |  | 13/03/2020 | 0 | 0.00 | 0.00 | 0.00 | 0.00 | 7.18 | 11 |
| 14 |  | 14/03/2020 | 0 | 0.00 | 0.00 | 0.60 | 0.00 | 7.83 | 12 |
| 15 |  | 15/03/2020 | 0 | 0.00 | 0.00 | 1.00 | 0.00 | 8.48 | 14 |
| 16 |  | 16/03/2020 | 0 | 0.00 | 0.00 | 1.36 | 0.00 | 9.13 | 16 |
| 17 |  | 17/03/2020 | 0 | 0.00 | 0.00 | 1.70 | 0.00 | 9.78 | 18 |
| 18 |  | 18/03/2020 | 1 | 0.33 | 3.00 | 2.08 | 0.48 | 10.42 | 21 |
| 19 |  | 19/03/2020 | 2 | 1.00 | 2.00 | 1.82 | 1.10 | 11.07 | 23 |
| 20 |  | 20/03/2020 | 3 | 1.67 | 1.80 | 1.72 | 1.74 | 11.72 | 25 |
| 21 |  | 21/03/2020 | 4 | 2.33 | 1.71 | 1.68 | 2.38 | 12.37 | 28 |
| 22 |  | 22/03/2020 | 7 | 3.67 | 1.91 | 1.70 | 4.13 | 13.02 | 31 |
| 23 |  | 23/03/2020 | 9 | 5.33 | 1.69 | 1.63 | 5.51 | 13.66 | 34 |
| 24 |  | 24/03/2020 | 9 | 6.00 | 1.50 | 1.62 | 5.56 | 14.31 | 37 |
| 25 |  | 25/03/2020 | 10 | 6.33 | 1.58 | 1.63 | 6.12 | 14.96 | 40 |
| 26 |  | 26/03/2020 | 15 | 8.33 | 1.80 | 1.65 | 9.10 | 15.61 | 43 |
| 27 |  | 27/03/2020 | 17 | 10.67 | 1.59 | 1.63 | 10.40 | 16.26 | 47 |
| 28 |  | 28/03/2020 | 20 | 12.33 | 1.62 | 1.63 | 12.29 | 16.90 | 50 |
| 29 |  | 29/03/2020 | 22 | 14.00 | 1.57 | 1.62 | 13.54 | 17.55 | 54 |
| 30 |  | 30/03/2020 | 27 | 16.33 | 1.65 | 1.64 | 16.50 | 18.20 | 58 |
| 31 | April | 01/04/2020 | 37 | 21.33 | 1.73 | 1.62 | 22.85 | 18.85 | 62 |
| 32 |  | 02/04/2020 | 40 | 25.67 | 1.56 | 1.59 | 25.17 | 19.49 | 66 |
| 33 |  | 03/04/2020 | 46 | 28.67 | 1.60 | 1.60 | 28.78 | 20.14 | 70 |
| 34 |  | 04/04/2020 | 55 | 33.67 | 1.63 | 1.58 | 34.79 | 20.79 | 75 |
| 35 |  | 05/04/2020 | 60 | 38.33 | 1.57 | 1.57 | 38.21 | 21.44 | 79 |
| 36 |  | 06/04/2020 | 67 | 42.33 | 1.58 | 1.57 | 42.62 | 22.09 | 84 |
| 37 |  | 07/04/2020 | 77 | 48.00 | 1.60 | 1.56 | 49.22 | 22.73 | 88 |
| 38 |  | 08/04/2020 | 79 | 52.00 | 1.52 | 1.55 | 51.08 | 23.38 | 93 |
| 39 |  | 09/04/2020 | 88 | 55.67 | 1.58 | 1.56 | 56.31 | 24.03 | 98 |
| 40 |  | 10/04/2020 | 97 | 61.67 | 1.57 | 1.56 | 62.24 | 24.68 | 103 |
| 41 |  | 11/04/2020 | 103 | 66.67 | 1.55 | 1.56 | 66.15 | 25.33 | 109 |
| 42 |  | 12/04/2020 | 105 | 69.33 | 1.51 | 1.56 | 67.17 | 25.97 | 114 |
| 43 |  | 13/04/2020 | 120 | 75.00 | 1.60 | 1.61 | 74.30 | 26.62 | 120 |
| 44 |  | 14/04/2020 | 130 | 83.33 | 1.56 | 1.61 | 80.71 | 27.27 | 125 |
| 45 |  | 15/04/2020 | 142 | 90.67 | 1.57 | 1.61 | 88.07 | 27.92 | 131 |
| 46 |  | 16/04/2020 | 157 | 99.67 | 1.58 | 1.63 | 96.57 | 28.56 | 137 |
| 47 |  | 17/04/2020 | 227 | 128.00 | 1.77 | 1.62 | 139.73 | 29.21 | 243 |
| 48 |  | 18/04/2020 | 252 | 159.67 | 1.58 | 1.58 | 159.23 | 29.86 | 249 |
| 49 |  | 19/04/2020 | 276 | 176.00 | 1.57 | 1.59 | 173.86 | 30.51 | 298 |
| 50 |  | 20/04/2020 | 330 | 202.00 | 1.63 | 1.59 | 207.45 | 31.16 | 362 |
| 51 |  | 21/04/2020 | 362 | 230.67 | 1.57 | 1.58 | 229.77 | 31.80 | 416 |
| 52 |  | 22/04/2020 | 394 | 252.00 | 1.56 | 1.57 | 250.61 | 32.45 | 471 |
| 53 |  | 23/04/2020 | 452 | 282.00 | 1.60 | 1.57 | 287.71 | 33.10 | 518 |
| 54 |  | 24/04/2020 | 506 | 319.33 | 1.58 | 1.56 | 324.89 | 33.75 | 559 |
| 55 |  | 25/04/2020 | 546 | 350.67 | 1.56 | 1.55 | 352.76 | 34.40 | 596 |
| 56 |  | 26/04/2020 | 586 | 377.33 | 1.55 | 1.55 | 378.26 | 35.04 | 603 |
| 57 |  | 27/04/2020 | 633 | 406.33 | 1.56 | 1.55 | 409.51 | 35.69 | 659 |
| 58 |  | 28/04/2020 | 663 | 432.00 | 1.53 | 1.56 | 425.99 | 36.34 | 698 |
| 59 |  | 29/04/2020 | 696 | 453.00 | 1.54 | 1.59 | 438.16 | 36.99 | 708 |
| 60 |  | 30/04/2020 | 758 | 484.67 | 1.56 | 1.59 | 477.14 | 37.64 | 783 |
| 61 | May | 01/05/2020 | 795 | 517.67 | 1.54 | 1.59 | 501.38 | 38.28 | 841 |
| 62 |  | 02/05/2020 | 922 | 572.33 | 1.61 | 1.59 | 579.70 | 38.93 | 949 |
| 63 |  | 03/05/2020 | 1198 | 706.67 | 1.70 | 1.58 | 759.43 | 39.58 | 1257 |
| 64 |  | 04/05/2020 | 1259 | 819.00 | 1.54 | 1.55 | 811.98 | 40.23 | 1295 |
| 65 |  | 05/05/2020 | 1344 | 867.67 | 1.55 | 1.55 | 865.74 | 40.87 | 1374 |
| 66 |  | 06/05/2020 | 1456 | 933.33 | 1.56 | 1.55 | 936.36 | 41.52 | 1482 |
| 67 |  | 07/05/2020 | 1548 | 1001.33 | 1.55 | 1.55 | 997.26 | 42.17 | 1591 |
| 68 |  | 08/05/2020 | 1678 | 1075.33 | 1.56 | 1.55 | 1081.99 | 42.82 | 1699 |
| 69 |  | 09/05/2020 | 1786 | 1154.67 | 1.55 | 1.55 | 1154.77 | 43.47 | 1788 |
| 70 |  | 10/05/2020 | 1939 | 1241.67 | 1.56 | 1.54 | 1256.75 | 44.11 | 1887 |
| 71 |  | 11/05/2020 | 2063 | 1334.00 | 1.55 | 1.54 | 1343.32 | 44.76 | 2124 |
| 72 |  | 12/05/2020 | 2173 | 1412.00 | 1.54 | 1.53 | 1417.20 | 45.41 | 2362 |
| 73 |  | 13/05/2020 | 2290 | 1487.67 | 1.54 | 1.53 | 1495.48 | 46.06 | 2456 |
| 74 |  | 14/05/2020 | 2377 | 1555.67 | 1.53 | 1.53 | 1552.08 | 46.71 | 2552 |
| 75 |  | 15/05/2020 | 2461 | 1612.67 | 1.53 | 1.53 | 1605.40 | 47.35 | 2648 |
| 76 |  | 16/05/2020 | 2576 | 1679.00 | 1.53 | 1.53 | 1678.43 | 48.00 | 2746 |
| 77 |  | 17/05/2020 | 2677 | 1751.00 | 1.53 | 1.53 | 1746.94 | 48.65 | 2845 |
| 78 |  | 18/05/2020 | 2825 | 1834.00 | 1.54 | 1.53 | 1843.00 | 49.30 | 2946 |
| 79 |  | 19/05/2020 | 2961 | 1928.67 | 1.54 | 1.53 | 1934.83 | 49.95 | 3047 |
| 80 |  | 20/05/2020 | 3103 | 2021.33 | 1.54 | 1.53 | 2025.36 | 50.59 | 3151 |
| 81 |  | 21/05/2020 | 3197 | 2100.00 | 1.52 | 1.53 | 2088.15 | 51.24 | 3255 |
| 82 |  | 22/05/2020 | 3332 | 2176.33 | 1.53 | 1.53 | 2172.18 | 51.89 | 3361 |
| 83 |  | 23/05/2020 | 3459 | 2263.67 | 1.53 | 1.53 | 2254.25 | 52.54 | 3468 |
| 84 |  | 24/05/2020 | 3667 | 2375.33 | 1.54 | 1.54 | 2380.17 | 53.18 | 3576 |
| 85 |  | 25/05/2020 | 3816 | 2494.33 | 1.53 | 1.54 | 2476.67 | 53.83 | 3885 |
| 86 |  | 26/05/2020 | 4009 | 2608.33 | 1.54 | 1.54 | 2595.88 | 54.48 | 4096 |
| 87 |  | 27/05/2020 | 4192 | 2733.67 | 1.53 | 1.55 | 2696.45 | 55.13 | 4265 |
| 88 |  | 28/05/2020 | 4536 | 2909.33 | 1.56 | 1.56 | 2911.62 | 56.42 | 4637 |
| 89 |  | 29/05/2020 | 4813 | 3116.33 | 1.54 | 1.55 | 3096.94 | 57.72 | 4953 |
| 90 |  | 30/05/2020 | 5130 | 3314.33 | 1.55 | 1.55 | 3302.27 | 58.37 | 5247 |
| 91 | June | 01/06/2020 | 5772 | 3634.00 | 1.59 | 1.54 | 3738.56 | 60.96 | 5847 |
| 92 |  | 02/06/2020 | 6168 | 3980.00 | 1.55 | 1.53 | 4023.99 | 63.55 | 6332 |
| 93 |  | 03/06/2020 | 6508 | 4225.33 | 1.54 | 1.53 | 4261.51 | 68.09 | 6536 |
| 94 |  | 04/06/2020 | 6876 | 4461.33 | 1.54 | 1.52 | 4521.83 | 72.62 | 6938 |
| 95 |  | 05/06/2020 | 7303 | 4584.00 | 1.50 | 1.50 | 4584.00 | 75.38 | 7354 |

## Data Availability

For any type of date, one can contact me in

https://orcid.org/0000-0002-5912-6287

